# Age-dependent differences in functional brain networks are atypical in Tourette syndrome

**DOI:** 10.1101/2020.04.06.20049817

**Authors:** Ashley N. Nielsen, Caterina Gratton, Soyoung Kim, Jessica A. Church, Kevin J. Black, Steven E. Petersen, Bradley L. Schlaggar, Deanna J. Greene

**Affiliations:** Institute for Innovations in Developmental Sciences, Northwestern University, Chicago, IL; Department of Medical Social Sciences, Northwestern University, Chicago, IL; Department of Psychology, Northwestern University, USA; Department of Neuroscience, Northwestern University, USA; Department of Neurology, Washington University School of Medicine, USA; Department of Psychiatry, Washington University School of Medicine, USA; Department of Psychology, The University of Texas at Austin, USA; Department of Neuroscience, Washington University School of Medicine, USA; Department of Radiology, Washington University School of Medicine, USA; Kennedy Krieger Institute, Baltimore, MD; Department of Neurology, Johns Hopkins University School of Medicine, Baltimore, MD; Department of Pediatrics, Johns Hopkins University School of Medicine, Baltimore, MD

**Keywords:** Tourette syndrome, development, functional connectivity, brain networks

## Abstract

Tourette syndrome (TS) is a neurodevelopmental disorder characterized by motor and vocal tics. TS is complex, with symptoms that involve sensory, motor, and top-down control processes and that fluctuate over the course of development. While many have studied atypical brain structure and function associated with TS, the neural substrates supporting the complex range and time-course of symptoms is largely understudied. Here, we used functional connectivity MRI to examine functional networks across the whole-brain in children and adults with TS. To investigate the functional neuroanatomy of childhood and adulthood TS, we separately considered the sets of connections within each functional network and those between each pair of functional networks. We tested whether developmental stage (child, adult), diagnosis (TS, control), or an interaction between these factors was present among these connections. We found that developmental changes for most functional networks in TS were unaltered (i.e., developmental differences in TS were similar to those in typically developing children and adults). However, there were several within-network and cross-network connections that exhibited either “divergent” or “attenuated” development in TS. Connections involving the somatomotor, cingulo-opercular, auditory, dorsal attention, and default mode networks diverged from typical development in TS, demonstrating enhanced functional connectivity in adulthood TS. In contrast, connections involving the basal ganglia, thalamus, cerebellum, auditory, visual, reward, and ventral attention networks showed attenuated developmental differences in TS. These results suggest that adulthood TS is characterized by increased functional connectivity among functional networks that support cognitive control and attention, which may be implicated in suppressing, producing, and attending to tics. In contrast, subcortical systems that have been implicated in the initiation and production of tics may be immature in adulthood TS. Jointly, our results reveal how several cortical and subcortical functional networks interact and differ across development in TS.

## Introduction

Tourette syndrome (TS) is a developmental neuropsychiatric disorder that affects 1-3% of children (Khalifa and Knorring, n.d.; Scahill *et al*., 2009; Cubo *et al*., 2011) and is characterized by motor and vocal tics (Leckman *et al*., 2014). Tics are brief, unwanted, repetitive movements or noises. Tics are often accompanied by a preceding perceived sensation of discomfort called a premonitory urge (Leckman *et al*., 1993); urges to tic can be suppressed, but only temporarily (Himle *et al*., 2007). Thus, TS is a complex disorder which affects multiple sensory, motor, and top-down control processes (Mink, 2001). Additionally, tic symptoms are not static and often fluctuate over the course of development. Tic onset typically occurs at age 5-7 years, with tic severity peaking during late childhood/early adolescence (10-12 years), and with marked improvement or even remission after adolescence and into adulthood (Erenberg *et al*., 1987; Leckman *et al*., 1998, Peterson *et al*., 2001*a*; Bloch *et al*., 2006; Hassan and Cavanna, 2012); However, symptom progression varies substantially across individuals, with a sizeable subgroup of patients (∼60%) experiencing moderate to severe tics that persist into adulthood (Leckman *et al*., 1998; Pappert *et al*., 2003). Taking into account the complexity of the nature and course of symptoms when studying the neural abnormalities in TS may provide better targets for diagnosis, treatments, and prognosis.

Many cortical and subcortical systems likely support the initiation, production, and suppression of tics and other symptoms associated with TS. One prominent theory in TS is that disruption of cortico-striato-thalamo-cortical loops leads to the production of tics; activity in the striatum propagates through these loops and leads to the disinhibition of unwanted motor plans and the production of tics (Mink, 2001). Several lines of research support this hypothesis as 1) disrupting activity in the basal ganglia produces tic-like movements (Alexander and DeLong, 1985; McCairn *et al*., 2009), 2) the basal ganglia, thalamus, motor cortex, and cerebellum are co-activated at the time of tic action in patients with TS (Bohlhalter *et al*., 2006; Wang *et al*., 2011; Neuner *et al*., 2014), and 3) reduced caudate volume and thinning of sensorimotor cortex have been observed in children and adults with TS (Peterson *et al*., 1993; Bloch *et al*., 2005, but see Greene *et al*., 2017). Concurrently, motor control and the suppression of unwanted movements are also atypical in TS and thought to be supported by a group of regions including frontal cortex. Regions in frontal cortex (and others) are active during the time preceding tics (premonitory urge) (Bohlhalter *et al*., 2006; Wang *et al*., 2011; Neuner *et al*., 2014) and during instructed eye blink suppression (Mazzone *et al*., 2010) in patients with TS. Control signals in frontal and other associated regions during non-motor tasks are also atypical in TS (Church *et al*., 2009) and thinning of frontal cortex has been observed in children with TS (Sowell *et al*., 2008). Less is known about the neurobiology supporting the initiation of tics; the frequency of tics can be affected by many environmental factors (e.g., stress, fatigue, diverted attention; for review see Conelea and Woods, 2008), which might suggest that the attention and sensory networks responsible for processing and orienting to external triggers might play an important role in the initiation of tics.

Many neuroimaging studies of TS treat it as a singular disorder, unchanging across development, by grouping together a wide range of patients or focusing on a single age cohort. However, as symptoms vary by age, there is evidence that differences in brain structure and function in TS also vary by age (Peterson *et al*., 2001*b*; Raz *et al*., 2009; Pépés *et al*., 2016). Considering whether brain differences in TS differ between childhood and adulthood enables one to determine whether an observed difference is necessary for the manifestation of tics over age. Further, given a context of typical development, one can determine whether brain differences observed in those with TS reflect atypically shifted development (e.g., accelerated or delayed maturation), potentially providing clues into etiology. Comparing the neural substrates supporting the initiation, production, and suppression of tics and other symptoms between children and adults with TS may reveal how maturation and/or experience with tics affect symptom course.

In combination, prior work suggests that many regions across the cortex and subcortex are implicated in TS, and any of these may exhibit changes over the course of development. Thus, studying the development of the network organization of the whole brain in TS is critical for a more complete understanding of the neural substrates of tic disorders. Functional brain networks can be examined using resting-state functional connectivity MRI: as fMRI signals from a pair of functionally related regions are often highly correlated even at rest (Biswal *et al*., 1995), measurements of “functional connectivity” have been used to identify collections of functionally related regions, or functional networks (Power *et al*., 2011; Yeo *et al*., 2011). Therefore, functional networks implicated in the array of TS symptoms can be rapidly and simultaneously assessed using resting-state functional connectivity.

Previously, we demonstrated that patterns of resting-state functional connectivity across the whole brain contain information that can distinguish individuals with TS from controls (Greene *et al*., 2016; Nielsen *et al*., 2020) and predict an individual’s maturity (Dosenbach *et al*., 2010; Nielsen *et al*., 2019, 2020). However, the specific functional networks that are altered in TS and how these connections differ in childhood vs. adulthood TS remains unknown. Here, we used a whole-brain network-level approach to investigate developmental differences in functional networks in TS compared to typical developmental differences observed in healthy controls. We separately considered sets of connections within each functional network and those between each pair of functional networks across the brain. We examined functional connectivity differences due to age (i.e., developmental stage: childhood, adulthood), diagnosis (TS, control), and their interaction.

## Materials and Methods

### Participants

A total of 172 individuals with TS, ages 7.3-35.0 years, were recruited from the Washington University School of Medicine Movement Disorders Center and the Tourette Association of America Missouri chapter. After quality control assessments of the neuroimaging data (see below) and to provide consistency with (Nielsen *et al*., 2020), 39 children (ages 7.6-13.1) and 39 adults (ages 18.4-35.0) with TS were included (Table 1). A group of 39 child and 39 adult control participants was selected from an extant database (n=487, ages 6.0–35.0 years, 206 males; recruited from the Washington University campus and surrounding community) and matched to the TS group on age, sex, IQ, handedness, and in-scanner movement (Table 1). Conditions commonly comorbid with TS (e.g., ADHD, OCD, anxiety) and medication use were not considered exclusionary for the TS group (Greene *et al*., 2016*a*) (Table S1) but were for the control group. All participants completed assessments of IQ, and TS participants completed additional assessments of symptom severity for TS, ADHD, and OCD (Supplement A). Adult participants and a parent or guardian for all child participants gave informed consent and all children assented to participation. The Washington University Human Research Protection Office approved all studies.

**Table 1.** Participant Characteristics.

| **Children** | **TS group** | **Control group** |
| --- | --- | --- |
| *N* | 39 | 39 |
| Male/Female | 31/8 | 30/9 |
| Age (Years) | 10.9 (1.6); 7.6-13.1 | 10.7 (1.5); 7.4-12.9 |
| Handedness (R/L) | 36/3 | 37/2 |
| IQ | 110 (12.3); 87-135 | 115 (12.1); 90-139 |
| Residual in-scanner movement (mean FD) | 0.11 (0.011); 0.092-0.14 | 0.11 (0.013); 0.067-0.13 |
| Amount of data ("good" frames) | 241.2 (69.4); 121-363 | 242.6 (88.2); 139-555 |
| YGTSS Total Tic Score | 17.5 (5.5); 0-37 | N/A |
| ADHD Rating Scale | 11.0 (8.2); 0-34 | N/A |
| CY-BOCS Score | 4.8 (5.5); 0-19 | N/A |

| Number on medications | 19 | 0 |
| --- | --- | --- |
| Number with comorbidities | 23 | 0 |
| **Adults** | **TS group** | **Control group** |
| *N* | 39 | 39 |
| Male/Female | 16/23 | 16/23 |
| Age (Years) | 25.9 (5.3); 18.4-35.0 | 25.9 (4.6); 18.1-34.2 |
| Handedness (R/L) | 36/3 | 36/3 |
| IQ | 119 (12.7); 83-139 | 119 (14.8); 83-145 |
| Residual in-scanner movement (mean FD) | 0.11 (0.017); 0.063-0.13 | 0.10 (0.012); 0.077-0.13 |
| Amount of data ("good" frames) | 349.7 (103.9); 153-573 | 311.3 (115.9); 170-668 |
| YGTSS Total Tic Score | 16.7 (8.3); 0-32 | N/A |
| ADHD Rating Scale | 11.9 (12.3); 0-44 | N/A |
| CY-BOCS Score | 6.8 (6.6); 0-24 | N/A |
| Number on medications | 18 | 0 |
| Number with comorbidities | 28 | 0 |

Where applicable values are displayed as Average (Standard Deviation); Range
FD = Frame-wise Displacement (in millimeters) (Power *et al.*, 2012a)
YGTSS = Yale Global Tic Severity Score (Total Tic Score) (Leckman *et al.*, 1989)
CY-BOCS Score = Children's Yale-Brown Obsessive-Compulsive Scale (Scahill *et al.*, 1997)

### Image Acquisition and Processing

Resting-state fMRI data were collected as participants viewed a centrally presented white crosshair on a black background. Participants were instructed to relax, look at the plus sign, and hold as still as possible. The duration and number of resting-state scans varied across participants (Supplement B). Imaging data were collected using a 3T Siemens Trio Scanner with a 12-channel Head Matrix Coil. Images were pre-processed to reduce artifacts (Shulman *et al*., 2010). Additional pre-processing steps were applied to the resting-state data to reduce spurious correlated variance unlikely related to neuronal activity. Stringent frame censoring (frame-wise displacement>0.2 mm) and nuisance regression (motion estimates, global signal, and individual ventricular and white matter signals) were used to reduce spurious individual or group differences in functional connectivity related to head movement in the scanner (Power *et al*., 2012*b*, 2014; Ciric *et al*., 2017). Participants with at least 5 minutes of low-motion data were included.

### Regions, Networks, and Blocks

For each participant, resting-state time-courses were extracted from a set of 300 regions of interest (ROIs) covering much of the cortex (Power *et al*., 2011), subcortex, and cerebellum (Figure 1; (Seitzman *et al*., 2020) available at https://greenelab.wustl.edu/data_software). Functional connectivity was measured as the correlation (Fisher z-transformed) between the resting-state time-courses for each pair of ROIs. Whole-brain functional connectivity among all 300 ROIs was determined for each of the four groups (control children, control adults, TS children, TS adults).

**Figure 1.**
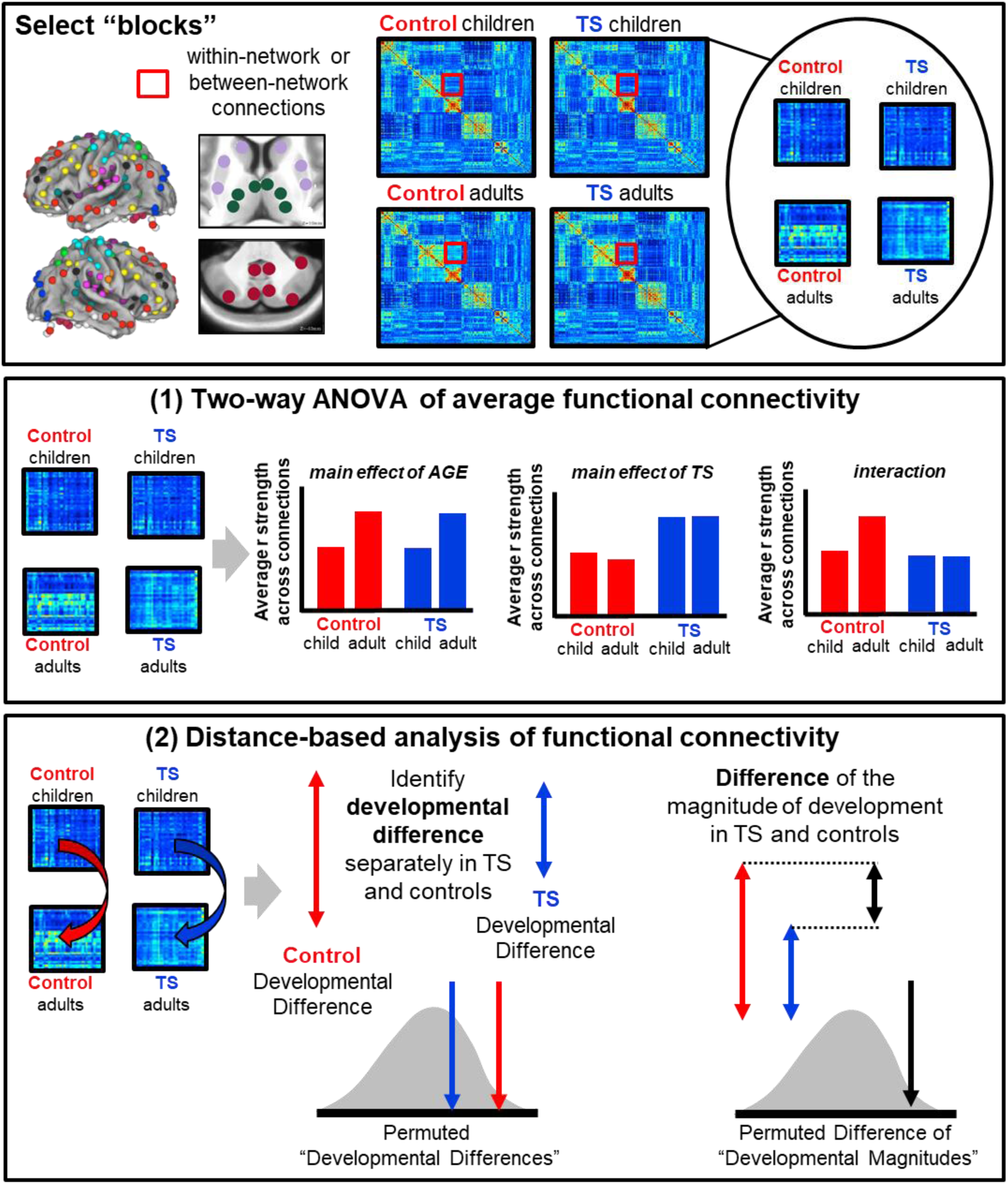
Overview of approaches to investigate the development of functional networks in Tourette syndrome.

Whole-brain functional connectivity has been shown to have modular organization such that different collections of ROIs assemble into separate functional networks thought to support different types of functions. These include processing networks (i.e., visual (VIS), auditory (AUD), and somatomotor (SM_body_, SM_face_) that interface with the external world, control networks (i.e., fronto-parietal (FP), cingulo-opercular (CO), salience (SAL), dorsal attention (DAN), and ventral attention (VAN)) that direct cognitive resources, and other association networks (i.e., default mode (DMN), parietal memory (PM), middle temporal lobe (MTL), and reward (RW)) that support internal associations. We examined regions of interest (ROIs) within each of these networks in the cortex, as well as ROIs within the basal ganglia (BG), thalamus (THAL), and cerebellum (CBL), which often link with multiple cortical networks (Choi *et al*., 2012; Greene *et al*., 2014; Marek *et al*., 2018).

To precisely describe developmental differences in functional networks across the brain, we separately considered “blocks” of connections *within* each functional network and those *between* pairs of functional networks. The term “block” comes from the block pattern observed when constructing correlation matrices describing functional connectivity among ROIs (see Figure 1). Within-network blocks represent the set of connections among a group of ROIs belonging to a single functional network (e.g., among DMN regions; on-diagonal portion of the correlation matrix). Cross-network blocks represent connections between two groups of ROIs from separate functional networks (e.g. between DMN regions and VIS regions; off-diagonal portion of the correlation matrix). Figure 1 provides an overview of our investigation of the influence of age (i.e., childhood vs. adulthood) and diagnosis (TS, control) on functional networks.

We used two approaches to reduce the dimensionality of each block for statistical group comparisons. First, we averaged the strength of functional connectivity from an entire block. While this is a straightforward dimensionality reduction approach, averaging across the connections in a block can obfuscate true effects if the block is not uniform. Averaging inherently assumes that (1) the strength and sign of the connections in a block are uniform and that (2) the developmental or diagnostic differences present between groups are uniform and in the same direction. Differences in functional connectivity that are non-uniform (e.g., half connections increase and half connections decrease) are not easily detected by differences in average connectivity. Thus, to complement this approach we used a second distance-based approach that computes the difference between groups separately for each connection in a block before being combined (Supplement E). Distance-based approaches are sensitive to non-uniform differences in functional connectivity across a block. However, distance-based approaches alone cannot distinguish (1) the direction of group differences (e.g., children > adults or children < adults) or (2) the relative position of group differences (e.g., overall delay in TS). A combination of distance-and average-based approaches helps to detect and describe differences between children and adults in the TS and control groups.

## Average-based analysis of functional connectivity across blocks

### Two-way ANOVA of average functional connectivity across blocks

As a first step, we performed a balanced two-way ANOVA to determine the effect of age, diagnosis, and the interaction between these factors on the average functional connectivity of each block from children and adults with and without TS. P-values were corrected for multiple comparisons across the total number of blocks tested (n = 136) using the FDR approach (requiring that P(FDR) < 0.05).

## Distance-based analysis of functional connectivity across blocks

### Distance-based identification of developmental differences in TS and controls

Next, the difference in functional connectivity between children and adults was determined for each block using Euclidean distance (see Formula S1), which maintains the relationship of specific region pairs and allows detection of complex developmental differences that might be non-uniform. We calculated developmental differences across all blocks in both the control and TS groups.

Permutation testing was used to determine whether the magnitude of the developmental differences in the control group or the TS group were greater than expected by chance. Children and adults in the control group and TS group were separately permuted (N = 1000 times) and randomly assigned into a “child” or “adult” group. Developmental differences were then calculated as described above. For each block, the true developmental difference in the control group or the TS group was contrasted with the distribution of permuted “developmental differences” to generate a P-statistic. P-values were corrected for multiple comparisons across the total number of tested blocks (n = 136) using the FDR approach (requiring that P(FDR) < 0.05).

### Distance-based comparison of developmental differences in TS and controls

We compared the magnitude of developmental differences in TS to the developmental differences in controls. Comparisons were limited to the blocks with significant developmental differences in either TS or controls. For these blocks, we calculated the difference between the magnitude (as defined by Euclidean distance) of developmental differences in TS and the magnitude of the developmental differences in controls.

Permutation testing was used to determine whether the difference in magnitude of developmental differences in each group (i.e., is the child vs. adult difference in each group significantly different from each other) was greater than expected by chance. Individuals in the control group and TS group were simultaneously, randomly permuted across age and diagnosis (N = 1000 times) and assigned into the “control child”, “control adult”, “TS child”, or “TS adult” group. Developmental differences were then calculated and compared as described above. For each tested block, the true difference in the magnitude of developmental differences in TS and controls was contrasted with the distribution of permuted differences to generate a P-statistic. P-values were corrected for multiple comparisons across the total number of tested blocks, using the FDR approach (requiring that P(FDR) < 0.05).

We also conducted further analyses to test whether the observed effects in TS could be attributed to possibly confounding variables, including co-morbid ADHD, tic severity, and medication status (Supplement F).

## Grouping Significant Results

To aid visualization and interpretation, we grouped blocks with similarly altered developmental differences in TS using average functional connectivity. For example, suppose two blocks exhibited a smaller developmental difference in TS compared to controls. Distance-based approaches alone cannot distinguish (1) the direction of the typical developmental differences (e.g., children > adults or children < adults) or (2) the direction of the atypical developmental differences in TS (e.g., accelerated in childhood TS, immature in adulthood TS, etc.). Using a data-driven modularity-based approach, we grouped blocks with similarly altered functional connectivity. For each block with significantly altered development in TS, we calculated the average functional connectivity across these connections for each subject. Then, we identified paired blocks in which the variability in average functional connectivity across individuals was similar (correlation between blocks > 0.3). Optimized modularity was then used to group linked sets of similarly altered blocks (Supplement G). Further, post-hoc t-tests were used to determine whether the atypical development observed across blocks stems from TS-related differences in the children and/or the adults with TS.

## Results

We found that many within-network and cross-network blocks were impacted by age and by diagnosis to different extents (Figure 2; two-way ANOVA). Numerous within-network (on-diagonal in Figure 2) and cross-network (off-diagonal in Figure 2) blocks exhibited a main effect of age in both TS and control groups (Figure 2A & D). Interestingly, select blocks, largely between control networks and the SM networks, exhibited a main effect of diagnosis in both children and adults (Figure 2B); however, these effects did not survive correction for multiple comparisons (Figure 2E). Further, an interaction between age and diagnosis (Figure 2C) was found in several blocks, including within the DMN, between the DAN and SM networks, and between the BG and the VIS network; some of these effects were significant after multiple comparisons correction (Figure 2F).

**Figure 2.**
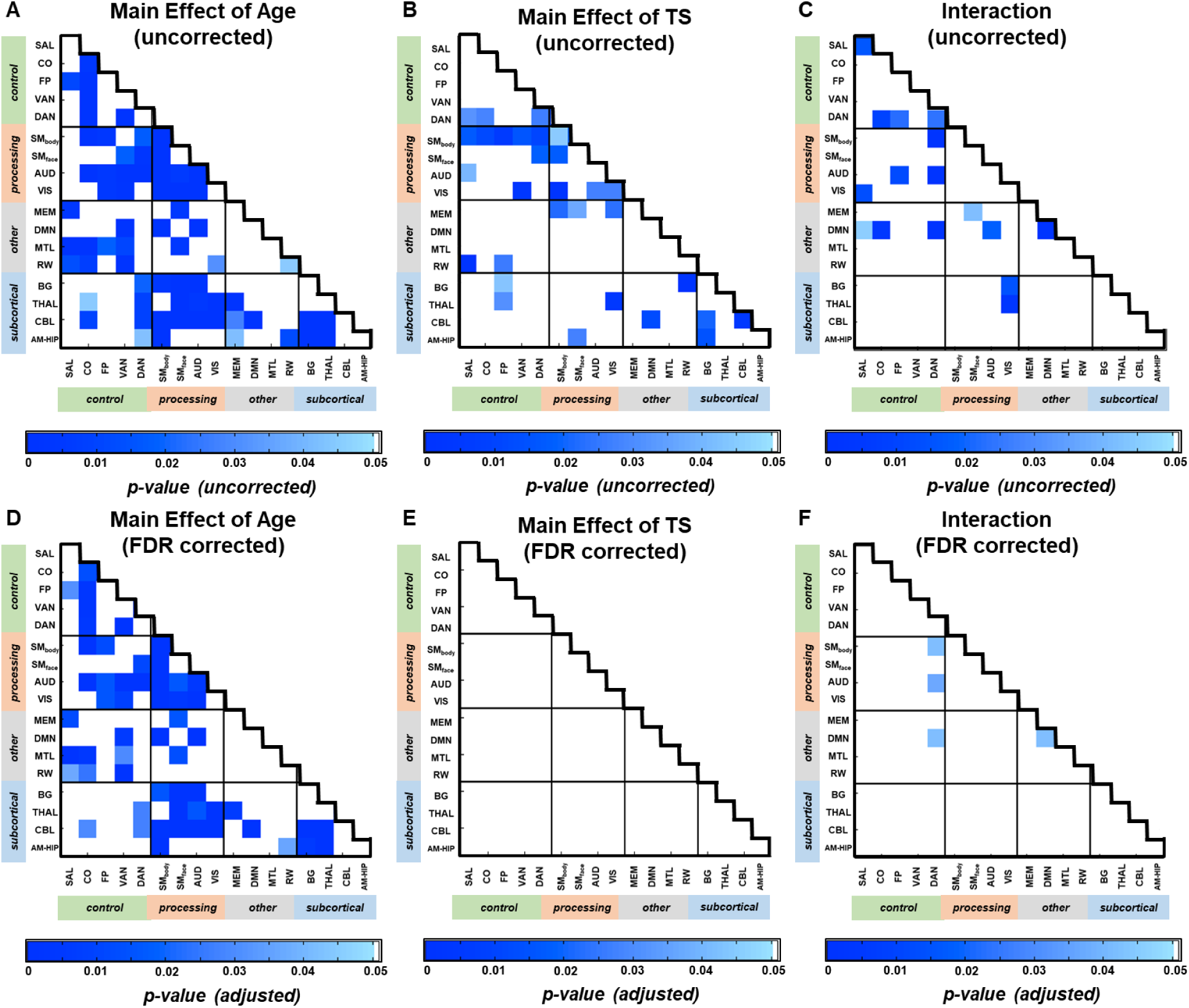
ANOVA of the average functional connectivity from each block. Within-network and cross-network blocks in which the average functional connectivity exhibits a main effect of age (A), diagnosis of TS (B), or an interaction between the two factors (C) before correcting for multiple comparisons. D-F depict the blocks with effects that survive after multiple comparisons correction.

When we compared functional connectivity among 300 regions spanning the whole brain between children and adults (i.e., developmental differences) with a distance-based approach, we also found many significant within-network and cross-network developmental differences in both the control group (Figure 3A) and the TS group (Figure 3B). Several within-network blocks (on-diagonal in Figure 3) differed between children and adults in only the control group (e.g. BG), only the TS group (e.g.,CO), or in both (e.g., SMbody). In addition, numerous cross-network blocks (off-diagonal in Figure 3) exhibited significant developmental differences in the control group (e.g. THAL-VIS), TS group (e.g., CO-DAN), or both (e.g., BG-SMbody). Critically, many of the developmental differences were observed in both groups; specifically, 4/10 within network blocks and 38/69 cross-network blocks shared significant developmental differences in TS and controls.

**Figure 3.**
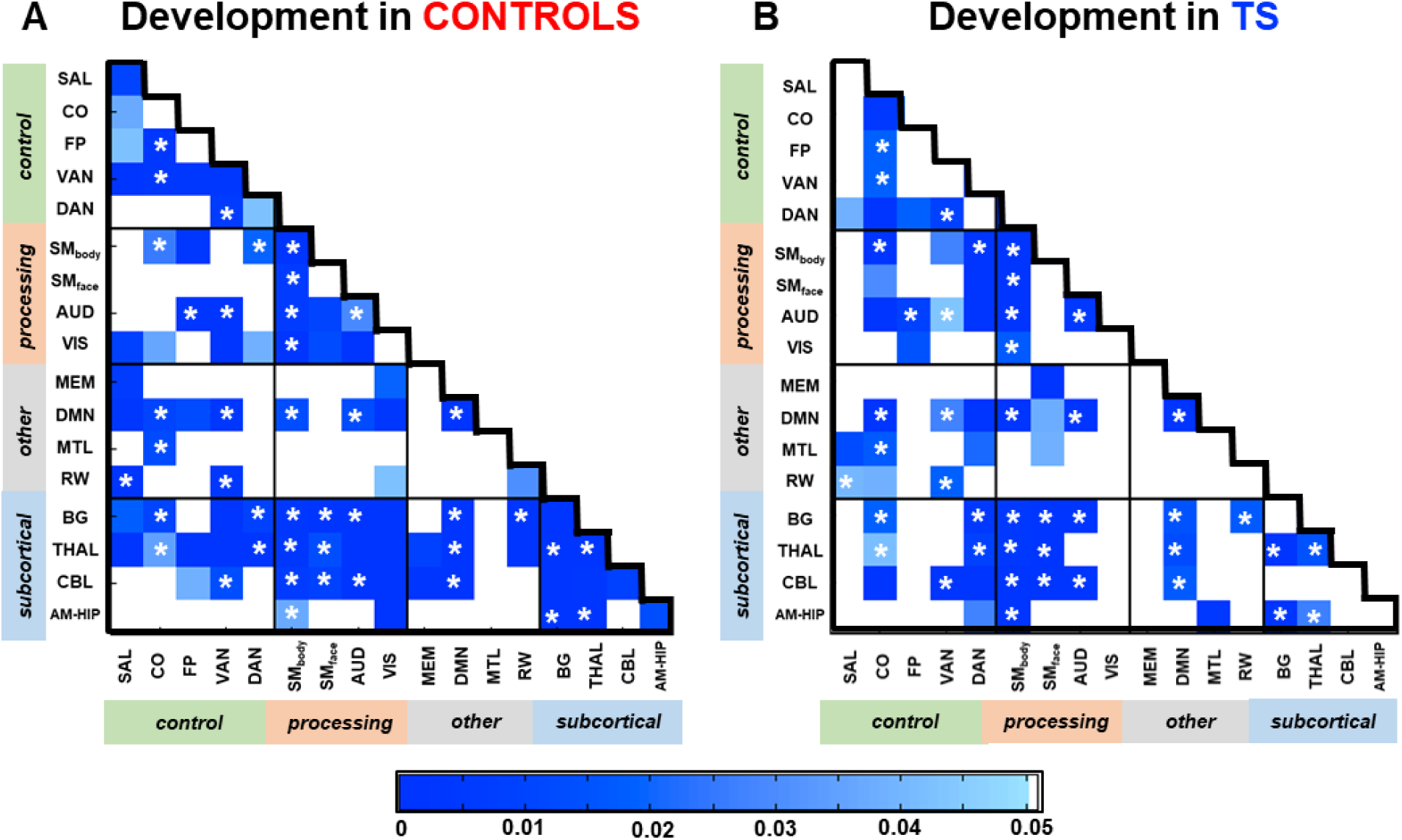
Developmental differences for each block in TS and controls. Within-network and cross-network blocks with significant developmental differences in the control group (A) and the TS group (B). The (*) indicates a developmental difference in both the control and TS groups.

When comparing developmental differences (child vs. adult) in functional networks in each group, we found specific blocks in which the magnitude of developmental differences was altered in TS (Figure S1). For several blocks (Figure 4A) and the block of connections within the CO network, the developmental differences observed in TS were *greater* than those observed in controls. Many (7/9) of these blocks with “divergent” development in TS did not exhibit significant developmental differences in the control group. In other cross-network blocks (Figure 4B) and the blocks of connections within the VAN, BG, and AMY/HIPP, the developmental differences observed in TS were *smaller* compared to those observed in controls. Many (15/17) of these blocks with “attenuated” development in TS actually did not exhibit significant developmental differences in the TS group.

**Figure 4.**
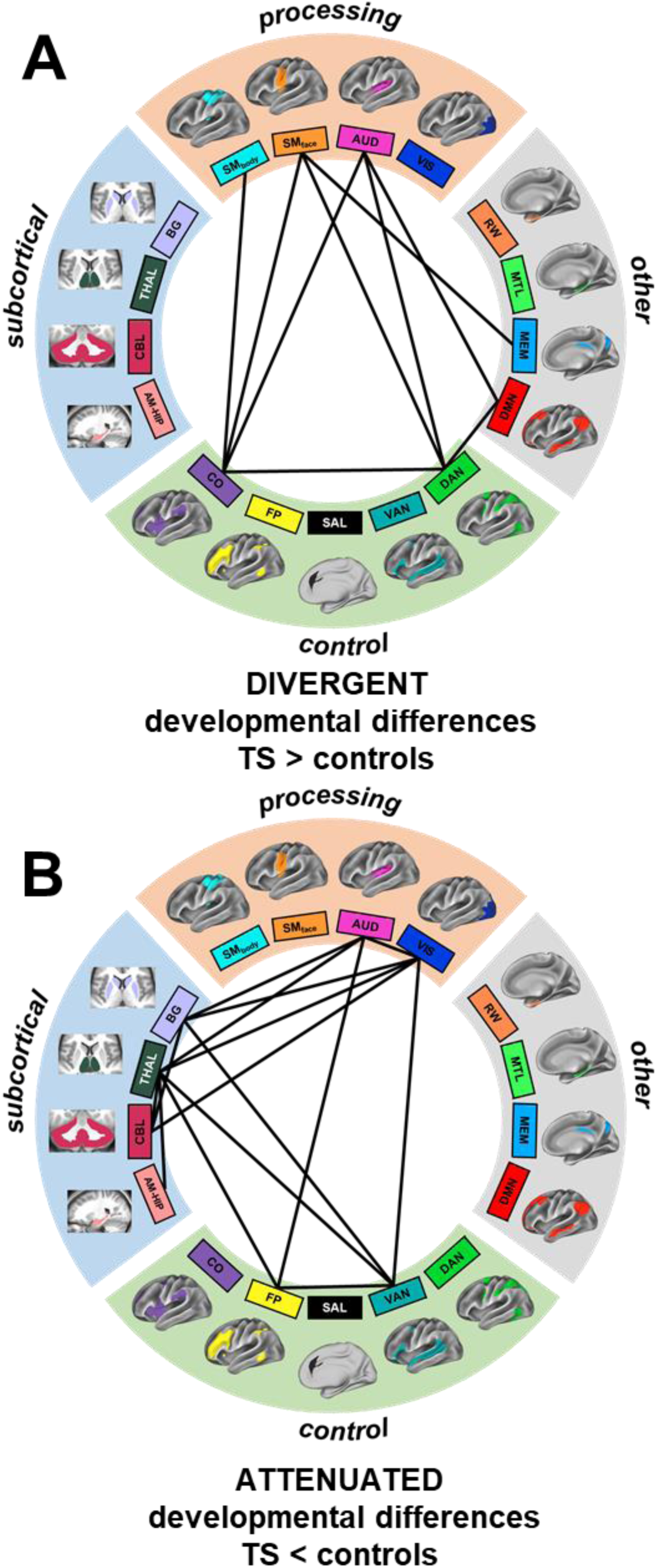
Functional network interactions exhibiting altered developmental differences in TS. Cross-network blocks with developmental differences that are (A) significantly greater in TS than in controls or (B) significantly greater in controls than in TS.

When we grouped blocks with average functional connectivity that was similarly altered in development in TS, four types of “atypical development” were identified (modularity = 0.59). These patterns were organized largely by type of alteration in TS (divergent vs. attenuated development) and by the direction of the developmental differences (children>adults or adults>children). The remaining blocks with idiosyncratically altered development in TS are described separately (Supplement H).

One group involved connections among the CO, DAN, SMbody, SMface, and AUD (Figure 5A). These blocks exhibited divergent age-dependent increases in functional connectivity in TS. Many of these blocks did not exhibit significant developmental differences in the controls (except CO-SM_body_). The connections among these control (CO and DAN) and sensorimotor networks (SM and AUD) were stronger in TS adults than in TS children (Figure 5B-C), and the average strength of many of these blocks was significantly greater in TS adults than in control adults (Table 2).

**Table 2.** Comparison of TS vs. controls in children and adults

|  | **Child TS vs. Controls**<br>FDR corrected p-values | **Adult TS vs. Controls**<br>FDR corrected p-values |
| --- | --- | --- |
| **Cluster #1** |  |  |
| CO – CO | 0.4256 | 0.0571 |
| CO - DAN | 0.7371 | 0.0034* |
| CO - SM<sub>body</sub> | 0.3342 | 0.0161* |
| DAN – SM<sub>face</sub> | 0.4002 | 0.0166* |
| CO – AUD | 0.9232 | 0.0273* |
| DAN – AUD | 0.1137 | 0.0006* |
| **Cluster #2** |  |  |
| DMN – DAN | 0.2196 | 0.0007* |
| DMN – AUD | 0.5840 | 0.0053* |
| **Cluster #3** |  |  |
| VAN – VIS | 0.2273 | 0.0043* |
| AUD – VIS | 0.7994 | 0.0083* |
| **Cluster #4** |  |  |
| BG – AUD | 0.9138 | 0.0319* |
| BG – VIS | 0.1323 | 0.0597 |
| THAL – AUD | 0.8639 | 0.2421 |
| THAL – VIS | 0.8471 | 0.0005* |
| THAL – RW | 0.3963 | 0.9021 |
| VIS – CBL | 0.8324 | 0.1451 |
| VAN – BG | 0.1287 | 0.2565 |
| VAN – THAL | 0.1250 | 0.4687 |
| **Remaining** |  |  |
| THAL – CBL | 0.7607 | 0.1133 |
| THAL – AM/HIP | 0.9007 | 0.5477 |
| AM/HIP-AM/HIP | 0.7190 | 0.0628* |
| SM<sub>face</sub> – MEM | 0.9708 | 0.0041* |
| VAN – VAN | 0.2035 | 0.3706 |
| SAL – DMN | 0.4746 | 0.0396* |
| BG – BG | 0.6652 | 0.0872 |
| FP – THAL | 0.2244 | 0.0680 |

**Figure 5.**
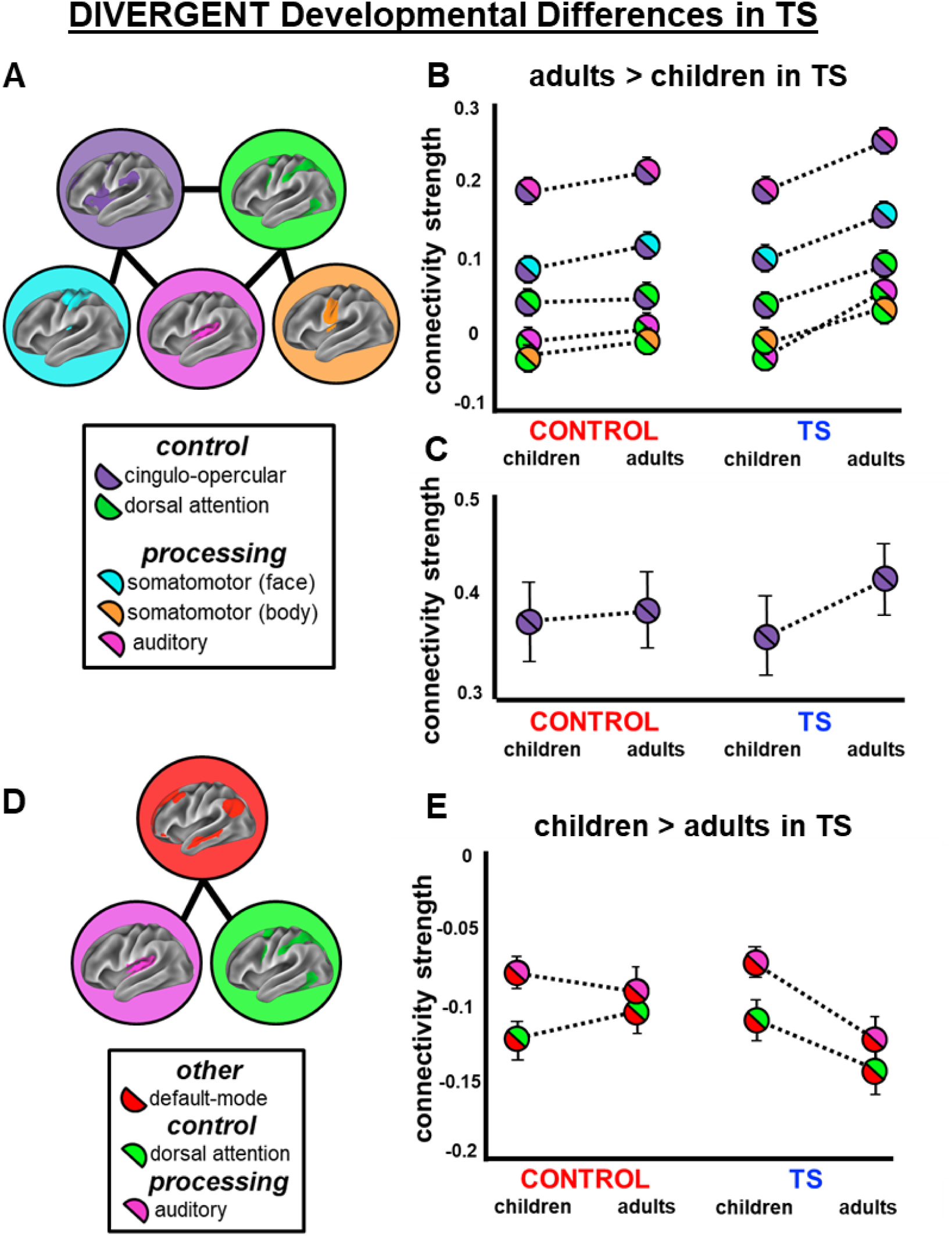
Functional network interactions exhibiting divergent developmental differences in TS. (A) Cluster of network interactions with similarly altered development in TS. Average functional connectivity of these cross-network (B) and within-network (C) blocks from (A) in the control children, control adults, TS children, and TS adults. (D) Cluster of network interactions with similarly altered development in TS. Average functional connectivity of these cross-network blocks (E) from (D) in the control children, control adults, TS children, and TS adults. Error bars are the stand error of the mean.

A second group involved connections between the DMN and the DAN and AUD networks (Figure 5D). These blocks exhibited divergent decreases in functional connectivity across development in TS. These blocks did not exhibit significant developmental differences in the controls, but were more strongly negative in the TS adults compared to the TS children (Figure 5E). The strength of the connections in these blocks was significantly more negative in adulthood TS compared to control adults (Table 2).

The third group involved connections between the VIS, VAN, and AUD networks (Figure 6A). These blocks exhibited attenuated increases in functional connectivity across development in TS. All of these blocks exhibited significant developmental differences in controls, but not in TS. These blocks were more strongly positive in adults compared to children in typical development, but not in TS (Figure 6B). The strength of connections between the VIS, VAN, and the AUD networks was significantly weaker in the TS adults than in the control adults (Table 2).

**Figure 6.**
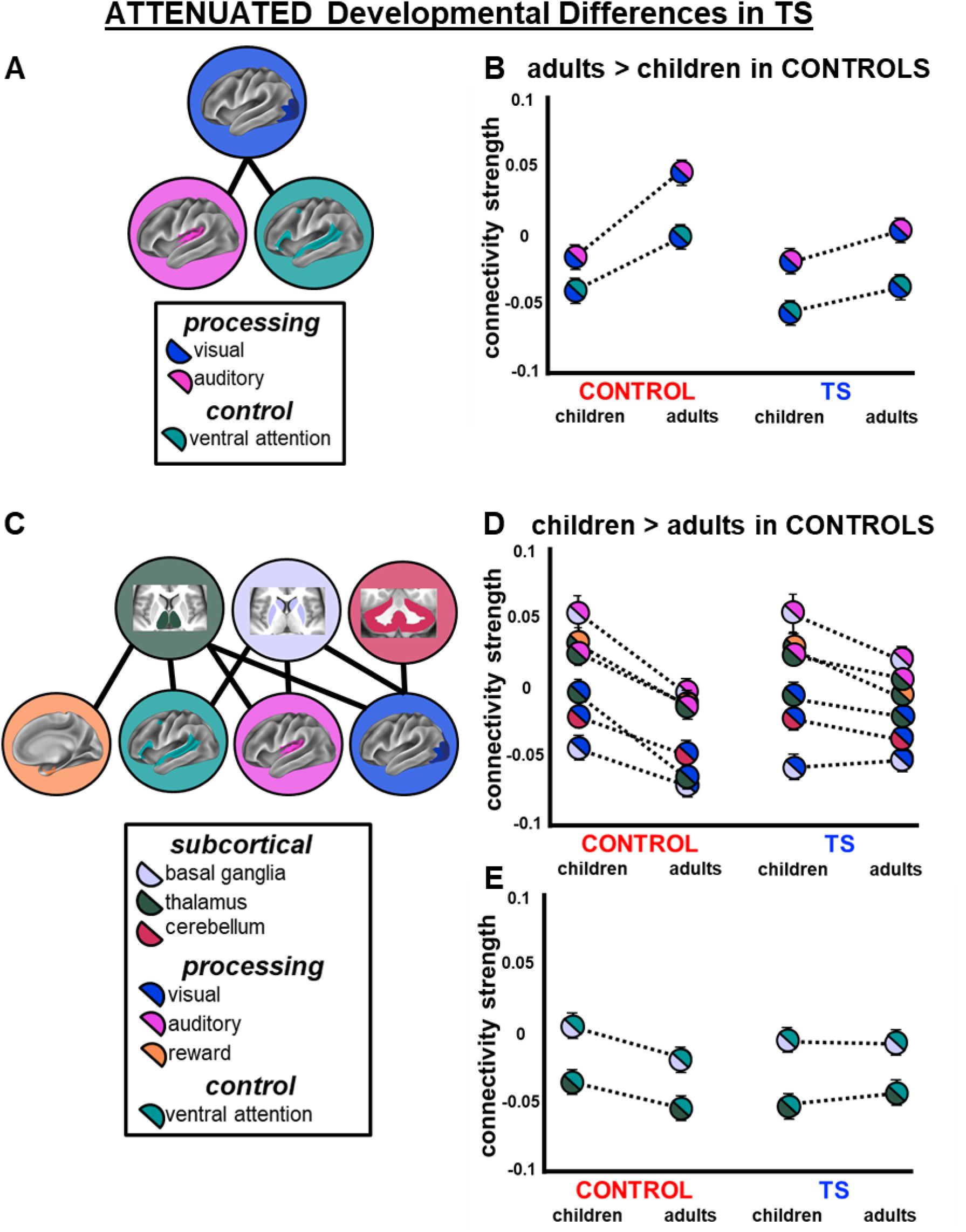
Functional network interactions exhibiting attenuated developmental differences in TS. (A) Cluster of network interactions with similarly altered development in TS. Average functional connectivity of these cross-network (B) blocks from (A) in the control children, control adults, TS children, and TS adults. (C) Cluster of network interactions with similarly altered development in TS. Average functional connectivity of these cross-network blocks (D & E) from (C) in the control children, control adults, TS children, and TS adults. Error bars are the stand error of the mean.

The fourth group involved connections between regions of the BG, THAL, and CBL and several sensorimotor (AUD, VIS), control (VAN), and other association (RW) networks (Figure 6C). These blocks exhibited attenuated decreases in functional connectivity across development in TS. Specifically, there were significant developmental differences in the controls, with stronger functional connectivity in the control children than in the control adults, yet the magnitude of these developmental differences was attenuated in TS. For the connections between the subcortical and processing functional networks (Figure 6D), the attenuation was driven by adulthood TS. The strength of the connections in these blocks was significantly less negative in TS adults than in control adults (Table 2). For the connections between the subcortical and VAN (Figure 6E), the attenuation was driven by initial negative connectivity in childhood TS. While not significant (Table 2, VAN-BG: p = 0.13, VAN-THAL: p = 0.12), the average strength of the connections in these blocks was more negative in TS children than in the control children.

Post-hoc analyses suggested that comorbid ADHD, tic severity, or the use of medications did not contribute to the observed altered developmental patterns in TS (Supplemental Table S2). Most differences between individuals with TS and controls were present even when individuals with ADHD, high tic severity, and current medications were removed.

## Discussion

In the present work, we applied a whole-brain network-level approach to functional connectivity MRI data in order to interrogate differences in functional brain organization in TS in two age groups: late childhood and early adulthood. We found that the organization of most functional networks in the TS groups was similar to that seen in typical development. However, there were several within-network and cross-network connections that exhibited either divergent or attenuated development in TS. Development of connections involving the SM, CO, AUD, DAN, and DMN networks diverged from typical development, with stronger functional connectivity in adulthood TS than in typically developing individuals and children with TS. Alternatively, typical developmental differences observed in connections involving the BG, THAL, CBL, AUD, VIS, RW, and VAN networks were reduced in TS. Together, these results suggest that different patterns of atypical development in TS involve different functional networks.

There is evidence from animal models (Alexander and DeLong, 1985; McCairn et al., 2009), human fMRI (Bohlhalter et al., 2006; Wang et al., 2011; Neuner et al., 2014), and transcranial magnetic stimulation (Ziemann et al., 1997) that subcortical structures (BG and THAL) and SM regions are involved in the production of tics in TS. Mink (2001) proposed that there is aberrant activity in cortico-striato-thalamo-cortical loops that leads to the disinhibition of unwanted motor plans and the production of tics. Interestingly, our results suggest that these cortical and subcortical regions are altered in different ways across development in TS. Specifically, we found stronger functional connectivity between sensorimotor and control networks in adulthood TS as compared to childhood TS and to typical development. In contrast, we found attenuated developmental differences in functional connectivity in the BG, THAL, and CBL in comparison to typical development, suggestive of immature connectivity in adulthood TS. This distinction shows that the developmental changes of the cortical and subcortical components of cortico-striato-thalamo-cortical circuitry differ, and that the developmental course of TS may be supported by a combination of developmental mechanisms.

We also found that functional connections involving several control networks (CO and DAN) and their connections with sensorimotor networks (SM_body_, SM_face_, AUD) were stronger in adulthood TS. In healthy controls, the CO network has been shown to be important for executive control, producing sustained signals that maintain goal-directed behaviors, detecting errors, conflict, and ambiguity (Dosenbach *et al*., 2007; Neta *et al*., 2014), while the DAN is important for directing cognitive resources, as in spatial attention (Corbetta and Shulman, 2002). In TS, regions in these control networks are also consistently activated during tic action and during the time preceding a tic (potentially related to the premonitory urge) (Bohlhalter *et al*., 2006). Activation of the CO network might reflect attempted tic suppression or the detection of an errant motor plan, while activation of the DAN might suggest that tics engage attentional resources. The stronger link between these control networks and sensorimotor networks in adulthood TS may be indicative of extended experience with tics and/or the premonitory urges associated with tics.

In contrast to the connections between sensorimotor and control networks, connections among subcortical structures (BG, THAL), sensory networks (AUD, VIS), and attention networks (VAN) showed attenuated age-related differences in TS compared to controls. This pattern is consistent with theories of incomplete/immature development in adulthood TS (Church *et al*., 2009). As VID and AUD networks process sensory inputs and the VAN is involved in reorienting attention in response to external stimuli (Corbetta and Shulman, 2002; Fox *et al*., 2006), connectivity between these networks in TS may be associated with environmental triggers (e.g., school vs. home setting) of tics (Conelea and Woods, 2008). Thus, disrupted development and immature segregation of these inputs to cortico-striatal-thalamo-cortical loops may be related to persistent tics in adulthood.

Given the potential involvement of cortico-striato-thalamo-cortical loops in TS, it is important to note that the present investigation may have oversimplified the functional organization of the basal ganglia, thalamus, and cerebellum. Separate cortico-striato-thalamo-cortical loops devoted to different functions (e.g, motor, control) involve specific regions within subcortical structures (Haber, 2003). These associations have been demonstrated in humans with resting-state functional connectivity, such that different regions within the subcortex and the cerebellum exhibit stronger functional connectivity with specific functional networks (Choi *et al*., 2012; Greene *et al*., 2014; Marek *et al*., 2018; Greene et al., 2020). In the present study, we treated each structure as a unit. However, it is possible that connections between specific regions of the subcortex/cerebellum and cortical functional networks develop differently in TS. Investigation of the functional organization of the basal ganglia, thalamus, and cerebellum at a finer scale may provide a more complete model of the neural substrates underlying the developmental course of TS.

While the present work has illuminated specific blocks of functional networks affected in TS and how these networks differ between children and adults, the source of these developmental differences is still undetermined. Some argue that childhood TS and adulthood TS are fundamentally different, given the commonly held belief that most patients with TS experience substantial symptom improvement or remission into adulthood (Eichele and Plessen, 2013). Therefore, by studying a sample of adults with current tics, we may have captured the subsample that does not experience remission. By contrast, any sample of children with TS will include a mixture of individuals whose tic symptoms will go on to improve and those whose tics will persist. However, there is evidence that remission is likely much rarer than previously estimated (10%, rather than 40%; Pappert *et al*., 2003b), and in our sample, many of the adults with TS reported improvement from childhood even if they did not report remission. Longitudinal data and studies of adults with remitted tics are necessary to determine to what extent the altered brain function in adulthood TS is a cause or consequence of prolonged symptom burden.

In addition, there are several possible sources of the disorder-related differences identified here. As an example, we identified atypically stronger functional connectivity between the CO network and the SM network. These strengthened connections might be a change in the brain that facilitates tics (e.g., atypically coordinated inhibitory control of motor function), a consequence of having chronic tics (experience with tics might produce maladaptive, neutral, or compensatory changes in the brain), or state differences between groups (subconscious tic suppression in the scanner in the TS group). Further investigation of the networks in TS with longitudinal developmental designs, links to experience with tics, and comparisons to studies of tic suppression is needed to shed light on the potential contribution of these various sources of disorder-related differences in functional connectivity.

Notably, our TS sample was highly representative of the TS population in that it was heterogeneous with respect to comorbid neuropsychiatric disorders, tic severity, and medication status (Freeman *et al*., 2000, Greene *et al*., 2016*a*). As brain network function can be affected by medications and other neuropsychiatric conditions (Mueller *et al*., n.d.), differences in functional connectivity observed in childhood and/or adulthood TS might have included medication-induced or comorbidity-related differences in brain function. Additionally, our child and adult samples differed with respect to sex; the children included more boys than girls, while the adults were more balanced. This difference reflects epidemiological data, as the sex imbalance (4:1 male:female) reported in childhood TS is attenuated in adulthood TS (Lichter and Finnegan, 2015). Nevertheless, we did not find evidence to suggest that these group differences contributed to our results. Still, future studies with larger samples will be useful for directly parsing the influence of medications, comorbidities, and sex on brain function in TS.

## Data Availability

Data may be made available upon request after the publication of this work.

## Acknowledgments

We thank Rebecca Coalson, Rebecca Lepore, Kelly McVey, Jonathan Koller, Annie Nguyen, Catherine Hoyt, Lindsey McIntyre, and Emily Bihun for assistance with data collection, the children and adults who participated in this study and their families, and Deanna Barch for her input on this manuscript. This project was supported by: Tourette Association of America fellowships (DJG, JAC), Tourette Association of America Neuroimaging Consortium pilot grant (KJB, BLS), Tourette Association of America research grant (DJG), NARSAD Young Investigator Award (DJG), NIH K01MH104592 (DJG), NIH R21MH091512 (BLS), NIH R01HD057076 (BLS), NIH R01NS046424 (SEP), NIH R21 NS091635 (BLS, KJB), NIH R01MH104030 (KJB, BLS), K12HD076224 (NUFD, Scholar of the Child Health Research Center at Washington University), NIH K23NS088590 (NUFD), NIH F32NS065649 (JAC), NIH F32NS092290 (CG), NIH F32NS656492, NIH K23DC006638, P50 MH071616, P60 DK020579-31 American Hearing Research Foundation, and The Simons Foundation Autism Research Initiative (“Brain circuitry in Simplex Autism,” SEP). Research reported in the publication was supported by the Eunice Kennedy Shriver National Institute of Child Health & Human Development of the National Institutes of Health under Award Number U54 HD087011 to the Intellectual and Developmental Disabilities Research Center at Washington University. The content is solely the responsibility of the authors and does not necessarily represent the official views of the National Institutes of Health.

## Supplemental Material

### A. Participants

A total of 78 children and adults with Tourette syndrome (TS) and 78 healthy control children and adults were included in the present study. All participants were native English speakers. All participants underwent a 2-scale brief assessment of IQ (WASI). For TS participants, the experimenter completed the following measures of “past week” symptom severity: Yale Global Tic Severity Score (Total Tic Score) (Leckman *et al*., 1989), Children’s Yale-Brown Obsessive Compulsive Scale (Scahill *et al*., 1997), and ADHD Rating Scale (Conners *et al*., 1998). All participants self-or parent-reported any history of neuropsychiatric diagnoses and current medications (Table S1). For the control participants, any history of neuropsychiatric or neurological diagnoses prohibited participation in the study.

**Table S1.**
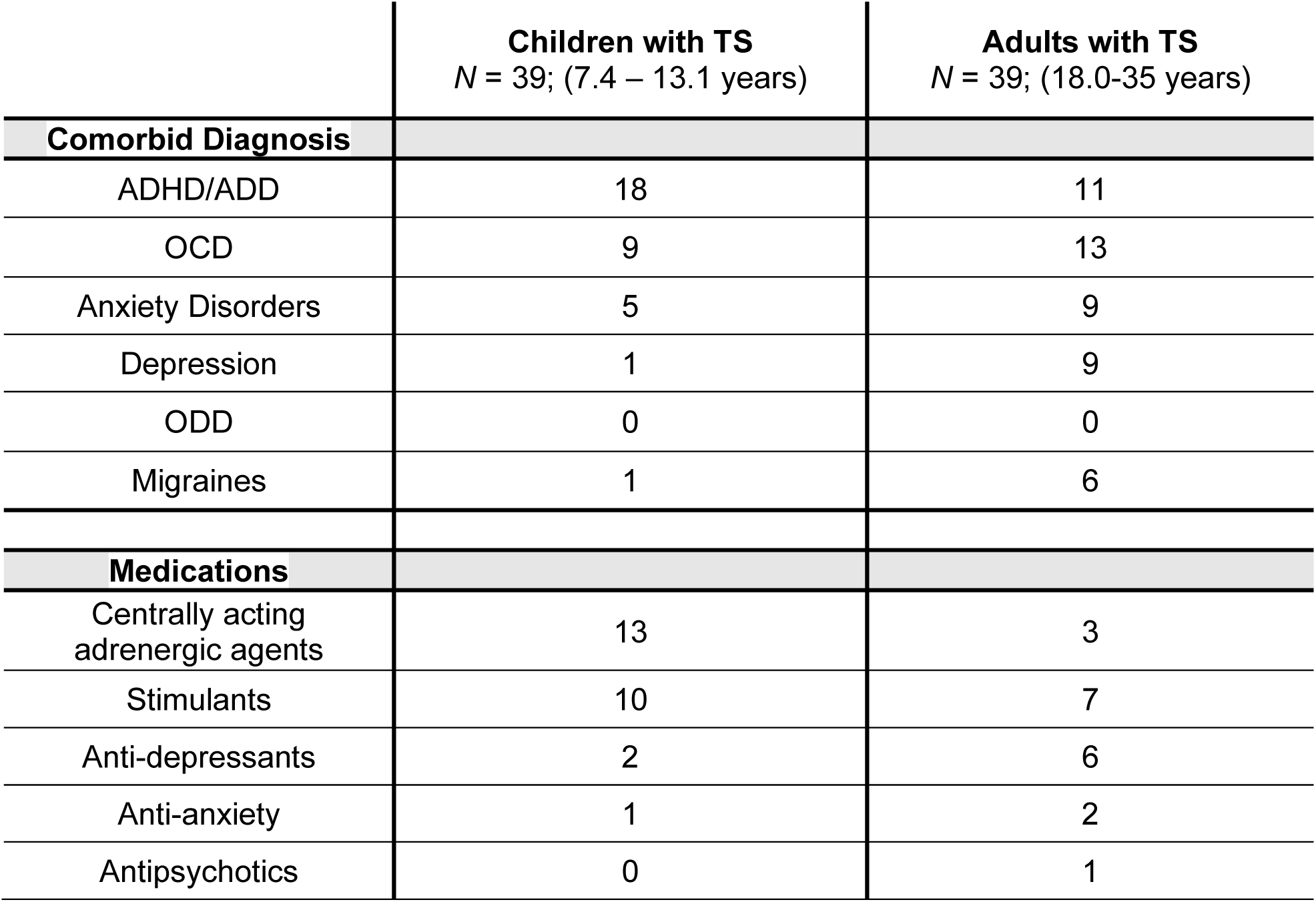
Comorbid diagnoses and current medications in participants with TS.

### B. Imaging Acquisition

Data were acquired on a Siemens 3T Trio scanner (Erlanger, Germany) with a Siemens 12-channel Head Matrix Coil. Each child was fitted with a thermoplastic mask fastened to the head coil to help stabilize head position. T1-weighted sagittal MP-RAGE structural images in the same anatomical plane as the BOLD images were obtained to improve alignment to an atlas (1 sequence acquisition for each of the 101 control participants (child, adolescent, and adult) and for 88 of the TS participants (child, adolescent, adult): slice time echo = 3.06 ms, TR = 2.4 s, inversion time = 1 s, flip angle = 8°, 176 slices, 1 × 1 × 1 mm voxels; 2 sequence acquisitions for each of the 13 remaining child and adolescent TS participants: slice time echo = 2.34 ms, TR = 2.2 s, inversion time = 1 s, flip angle = 7°, 160 slices, 1 × 1 × 1 mm voxels). Functional images were acquired using a BOLD contrast-sensitive echo-planar sequence (TE = 27 ms, flip angle = 90°, in-plane resolution 4×4 mm; volume TR = 2.5 s). Whole-brain coverage was obtained with 32 contiguous interleaved 4 mm axial slices. Steady-state magnetization was assumed after 4 volumes. For most participants, 2-4 resting state scans lasting 5-5.5 min each were acquired, but the duration of each scan ranged from 3.2 minutes to 30 minutes. In the TS group, 388 ± 61.5 (range 264-528) total functional volumes were acquired, and in the control group, 372 ± 130 (range 260-724) total functional volumes were acquired.

### C. Imaging preprocessing

Functional images from each participant were preprocessed to reduce artifacts (Shulman et al. 2010). These steps included: (i) temporal sinc interpolation of all slices to the temporal midpoint of the first slice, accounting for differences in the acquisition time of each individual slice, (ii) correction for head movement within and across runs, and (iii) intensity normalization of the functional data was computed for each individual via the MP-RAGE T1-weighted scans. Each run was then resampled in atlas space on an isotropic 3 mm grid combining movement correction and atlas transformation in a single interpolation. The target atlas was created from thirteen 7-9 year old children and twelve 21-30 year old adults using validated methods (Black et al. 2004). The atlas was constructed to conform to the Talairach atlas space.

### D. Functional Connectivity Preprocessing

Several additional pre-processing steps were applied to reduce spurious variance unlikely to reflect neuronal activity (Fox et al. 2009). These functional connectivity pre-processing steps included: (i) demeaning and detrending each run, (ii) multiple regression of nuisance variables,(iii) frame censoring (discussed below) and interpolation of data within each run, (iv) temporal band-pass filtering (0.009 Hz < f < 0.08 Hz), and (v) spatial smoothing (6 mm full width at half maximum). Nuisance variables included motion regressors (e.g. original motion estimates, motion derivatives, and Volterra expansion of motion estimates), an average of the signal across the whole brain (global signal), individualized ventricular and white matter signals, and the derivatives of these signals.

We applied a procedure determined and validated to best reduce artifacts related to head motion (Power et al. 2014; Ciric et al. 2017). Specifically, frame-by-frame head displacement (FD) was calculated from preprocessing realignment estimates, and frames with FD > 0.2 mm were removed. An FD threshold of 0.2 mm was chosen because it best reduced the distance-dependence related to individual differences in head motion (mean FD) in this developmental dataset, as assessed using procedures from Power et al. (2012) and Ciric et al. (2017). Data were considered usable only in contiguous sets of at least 3 frames with FD < 0.2 and a minimum of 30 frames within a functional run. Motion-contaminated frames were censored from the continuous, processed resting-state time series before computing resting-state correlations. Notably, the global signal was included as a nuisance regressor (mentioned above) in order to further reduce global, motion-related spikes in BOLD data (Power et al. 2014; Ciric et al. 2017) and reduce patterns of spurious functional connectivity that might be utilized for prediction with machine learning (Nielsen et al. 2018).

### E. Euclidean Distance Formula

Euclidean distance was used measure the difference in functional connectivity between children and adults and between TS and controls. This measure reduces the dimensionality of a set of connections in order to determine whether function connectivity differed between groups. For a set of functional connections the difference between two groups, children and adults, was calculated using Formula S1.

**Formula S1. Euclidean Distance Formula**

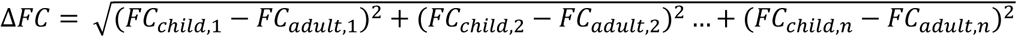

FC: functional connection

N: number of connections

Child: average functional connectivity across child group

Adult: average functional connectivity across adult group

### F. Effects of co-morbid ADHD, tic severity, medications on functional connectivity in TS

Many of the patients with TS had comorbid diagnoses of the other neuropsychiatric disorders and/or were taking medications (see Table S1). In addition, tic severity varied within the children and adults with TS. We investigated whether these factors contributed to the results observed in the main text.

Patients with TS in our sample were most commonly also diagnosed with ADHD. We examined whether TS-related differences in functional connectivity were present in individuals with and without an additional diagnosis of ADHD in children and adults separately. Next, we split our TS sample into two groups with low tic severity and high tic severity according to YGTSS. We then examined whether TS-related differences in functional connectivity were present in individuals with less severe and more severe tic symptoms in children and adults separately. Finally, we examined whether TS-related differences in functional connectivity were present in individuals that were and were not taking medications. Comparisons were limited to blocks with significant developmental differences in TS and are reported in Table S2.

**Table S2.** TS-related differences in functional connectivity before and after removing individuals with ADHD, high tic severity, and current medications.

| **Children with TS vs. Controls** | Total sample | w/ ADHD | w/o ADHD | High tic severity | Low tic severity | w/ current medications | w/o current medications |
| --- | --- | --- | --- | --- | --- | --- | --- |
| CO-CO | 0.426 | 0.455 | 0.549 | 0.845 | 0.274 | 0.707 | 0.308 |
| CO-DAN | 0.737 | 0.385 | 0.991 | 0.431 | 0.879 | 0.434 | 0.820 |
| CO-SMbody | 0.334 | 0.335 | 0.315 | 0.410 | 0.253 | 0.068 | 0.915 |
| DAN-SMface | 0.400 | 0.054 | 0.169 | 0.131 | 0.075 | 0.016 | 0.309 |
| CO-AUD | 0.923 | 0.958 | 0.485 | 0.838 | 0.597 | 0.569 | 0.873 |
| DAN-AUD | 0.114 | 0.820 | 0.487 | 0.765 | 0.338 | 0.786 | 0.776 |
| SMface-PM | 0.971 | 0.970 | 0.981 | 0.553 | 0.511 | 0.734 | 0.781 |
| DAN-DMN | 0.220 | 0.132 | 0.149 | 0.314 | **0.047\*** | 0.177 | 0.109 |
| AUD-DMN | 0.584 | 0.847 | 0.850 | 0.854 | 0.539 | 0.593 | 0.862 |
| VAN-VAN | 0.204 | 0.594 | 0.130 | 0.564 | 0.109 | 0.265 | 0.346 |
| VAN-VIS | 0.227 | 0.474 | 0.293 | 0.319 | 0.443 | 0.649 | 0.188 |
| AUD-VIS | 0.799 | 0.629 | 0.462 | 0.858 | 0.605 | 0.993 | 0.807 |
| SAL-DMN | 0.475 | 0.278 | 0.439 | 0.448 | 0.251 | 0.247 | 0.492 |
| VAN-BG | 0.129 | 0.283 | 0.085 | 0.045 | 0.489 | 0.065 | 0.342 |
| AUD-BG | 0.914 | 0.059 | 0.647 | 0.112 | 0.563 | 0.102 | 0.616 |
| VIS-BG | 0.132 | 0.297 | 0.438 | 0.735 | 0.176 | 0.996 | 0.106 |
| BG-BG | 0.665 | 0.450 | 0.217 | 0.906 | 0.469 | 0.499 | 0.240 |
| FP-THAL | 0.224 | 0.207 | **0.032\*** | 0.220 | **0.026\*** | 0.039 | 0.176 |
| VAN-THAL | 0.125 | **0.027\*** | 0.445 | **0.034\*** | 0.381 | 0.152 | 0.108 |
| AUD-THAL | 0.864 | **0.001\*** | 0.821 | **0.032\*** | 0.592 | 0.099 | 0.273 |
| VIS-THAL | 0.847 | 0.736 | 0.304 | 0.812 | 0.231 | 0.832 | 0.077 |
| RW-THAL | 0.396 | 0.923 | 0.383 | 0.248 | 0.931 | 0.748 | 0.193 |
| VIS-CBL | 0.832 | 0.357 | 0.277 | 0.122 | 0.727 | 0.141 | 0.559 |
| THAL-CBL | 0.761 | 0.656 | 0.778 | 0.666 | 0.765 | 0.895 | 0.567 |
| THAL-AMY-HIP | 0.901 | **0.001\*** | 0.147 | **0.008\*** | 0.060 | **0.039\*** | **0.013\*** |
| AMY/HIP-AMY/HIP | 0.719 | 0.488 | 0.206 | 0.902 | 0.611 | 0.644 | 0.315 |
| **Adults with TS vs. Controls** | Total sample | w/ ADHD | w/o ADHD | High tic severity | Low tic severity | w/ current medications | w/o current medications |
| CO-CO | 0.057 | 0.276 | 0.063 | 0.068 | 0.208 | **0.020\*** | 0.466 |
| CO-DAN | **0.003\*** | **0.021\*** | **0.001\*** | **0.000\*** | **0.026\*** | **0.002\*** | **0.005\*** |
| CO-SMbody | **0.016\*** | **0.044\*** | **0.015\*** | **0.008\*** | 0.070 | **0.007\*** | 0.081 |
| DAN-SMface | **0.017\*** | 0.050 | **0.009\*** | **0.006\*** | 0.057 | **0.026\*** | **0.015\*** |
| CO-AUD | **0.027\*** | 0.083 | **0.027\*** | 0.189 | **0.005\*** | 0.051 | **0.038\*** |
| DAN-AUD | **0.001\*** | 0.067 | **0.000\*** | **0.002\*** | **0.002\*** | **0.001\*** | **0.006\*** |
| SMface-PM | **0.004\*** | **0.017\*** | 0.058 | 0.157 | **0.007\*** | **0.031\*** | 0.077 |
| DAN-DMN | **0.001\*** | **0.023\*** | **0.004\*** | **0.000\*** | 0.097 | **0.000\*** | 0.083 |
| AUD-DMN | **0.005\*** | **0.018\*** | **0.006\*** | **0.023\*** | **0.003\*** | **0.014\*** | **0.007\*** |
| VAN-VAN | 0.371 | **0.023\*** | 0.786 | 0.563 | 0.380 | 0.493 | 0.439 |
| VAN-VIS | **0.004\*** | **0.045\*** | **0.024\*** | **0.003\*** | 0.139 | **0.008\*** | 0.067 |
| AUD-VIS | **0.008\*** | 0.170 | **0.040\*** | **0.001\*** | 0.523 | **0.030\*** | 0.154 |
| SAL-DMN | **0.040\*** | 0.728 | **0.047\*** | 0.419 | 0.095 | 0.062 | 0.481 |
| VAN-BG | 0.257 | 0.079 | 0.878 | 0.370 | 0.537 | 0.072 | 0.997 |
| AUD-BG | **0.032\*** | **0.009\*** | 0.620 | 0.078 | 0.431 | **0.019\*** | 0.569 |
| VIS-BG | 0.060 | 0.375 | **0.000\*** | **0.003\*** | **0.028\*** | **0.033\*** | **0.002\*** |
| BG-BG | 0.087 | 0.151 | 0.167 | **0.027\*** | 0.423 | **0.003\*** | 0.918 |
| FP-THAL | 0.068 | **0.042\*** | **0.044\*** | **0.009\*** | 0.153 | 0.214 | **0.009\*** |
| VAN-THAL | 0.469 | 0.069 | 0.714 | 0.394 | 0.744 | 0.382 | 0.926 |
| AUD-THAL | 0.242 | **0.028\*** | 0.510 | 0.361 | 0.523 | 0.179 | 0.449 |
| VIS-THAL | **0.001\*** | **0.033\*** | **0.000\*** | **0.000\*** | **0.001\*** | **0.010\*** | **0.000\*** |
| RW-THAL | 0.902 | 0.748 | 0.848 | 0.770 | 0.782 | 0.355 | 0.345 |
| VIS-CBL | 0.145 | 0.303 | 0.170 | **0.031\*** | 0.759 | 0.068 | 0.459 |
| THAL-CBL | 0.113 | 0.146 | 0.274 | 0.187 | 0.369 | **0.012\*** | 0.973 |
| THAL-AMY-HIP | 0.548 | 0.244 | **0.036\*** | 0.190 | **0.023\*** | 0.132 | 0.060 |
| AMY/HIP-AMY/HIP | 0.063 | **0.032\*** | 0.180 | 0.176 | 0.080 | **0.004\*** | 0.647 |

### G. Optimized Modularity to group blocks with similarly altered development in TS

Modularity optimization was used to group blocks with similarly altered development of functional connectivity in TS. First, we averaged the functional connectivity across each set of functional connections for each individual. Then we calculated how correlated this individual variability in average functional connectivity was between pairs of different blocks. We tested several thresholds (r = 0.1 – 0.5, increments of 0.05) to define “similarly altered” pairs of blocks. We calculated the modularity, a graph theoretical measure, of each network of similarly altered connections (Newman, 2006). Figure S1 depicts the modularity across these thresholds. We chose r = 0.3 in order to maximize modularity and retain more blocks.

**Figure S1.**
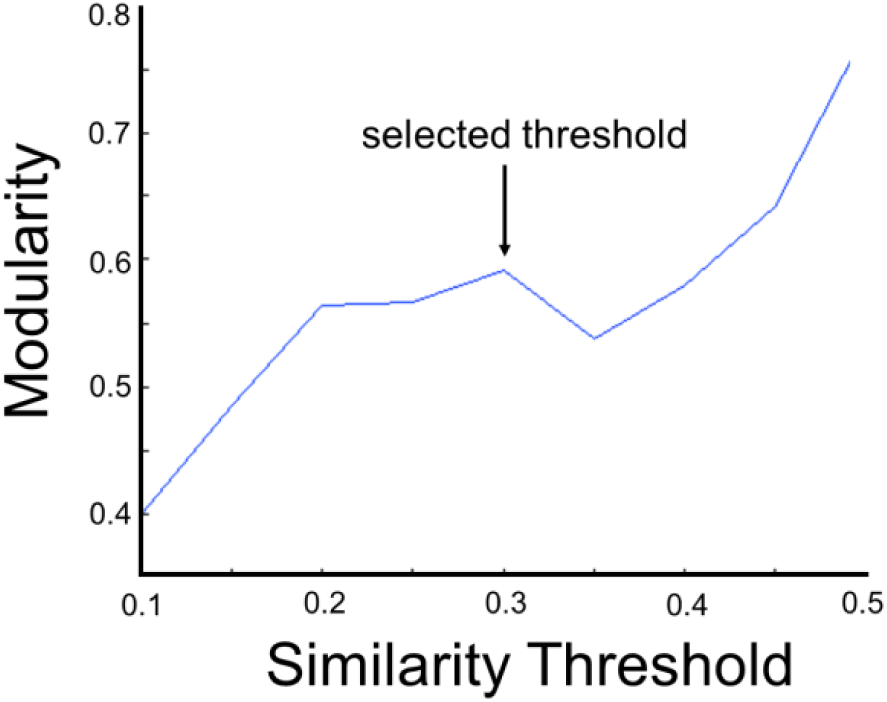
Modularity of the similarity across all individuals between blocks at difference thresholds.

### H. Divergent and attenuated developmental differences in TS

In the main text, we found that within-network and cross-network blocks with divergent and attenuated developmental differences in TS. Figure S2 depicts the FDR adjusted p-values that describe these effects.

**Figure S2.**
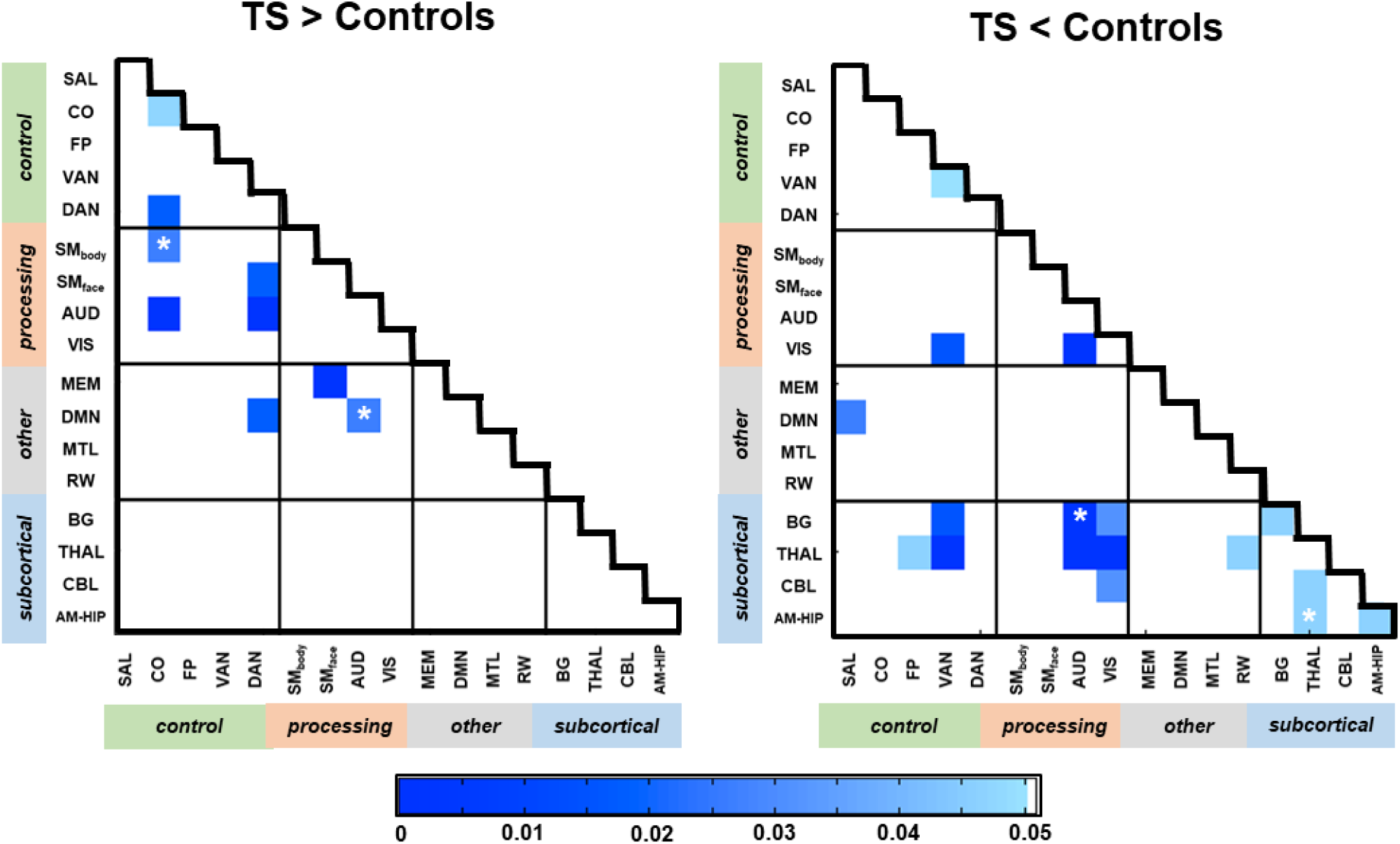
Blocks of within-network and cross-network functional connections in which the magnitude of developmental differences significantly differs in the control and TS groups. The left panel depicts blocks with divergent developmental differences that are greater in TS than in controls. The right panel depicts blocks attenuated developmental differences that are smaller in TS than in controls. The (*) indicates a significant developmental difference in both the control and TS groups. P-values are FDR adjusted.

### I. Remaining blocks with altered development in TS

Some blocks with significantly altered development in TS could not be grouped with other blocks. The average functional connectivity of these blocks across the children and adults with and without TS are shown in Figure S3.

**Figure S3.**
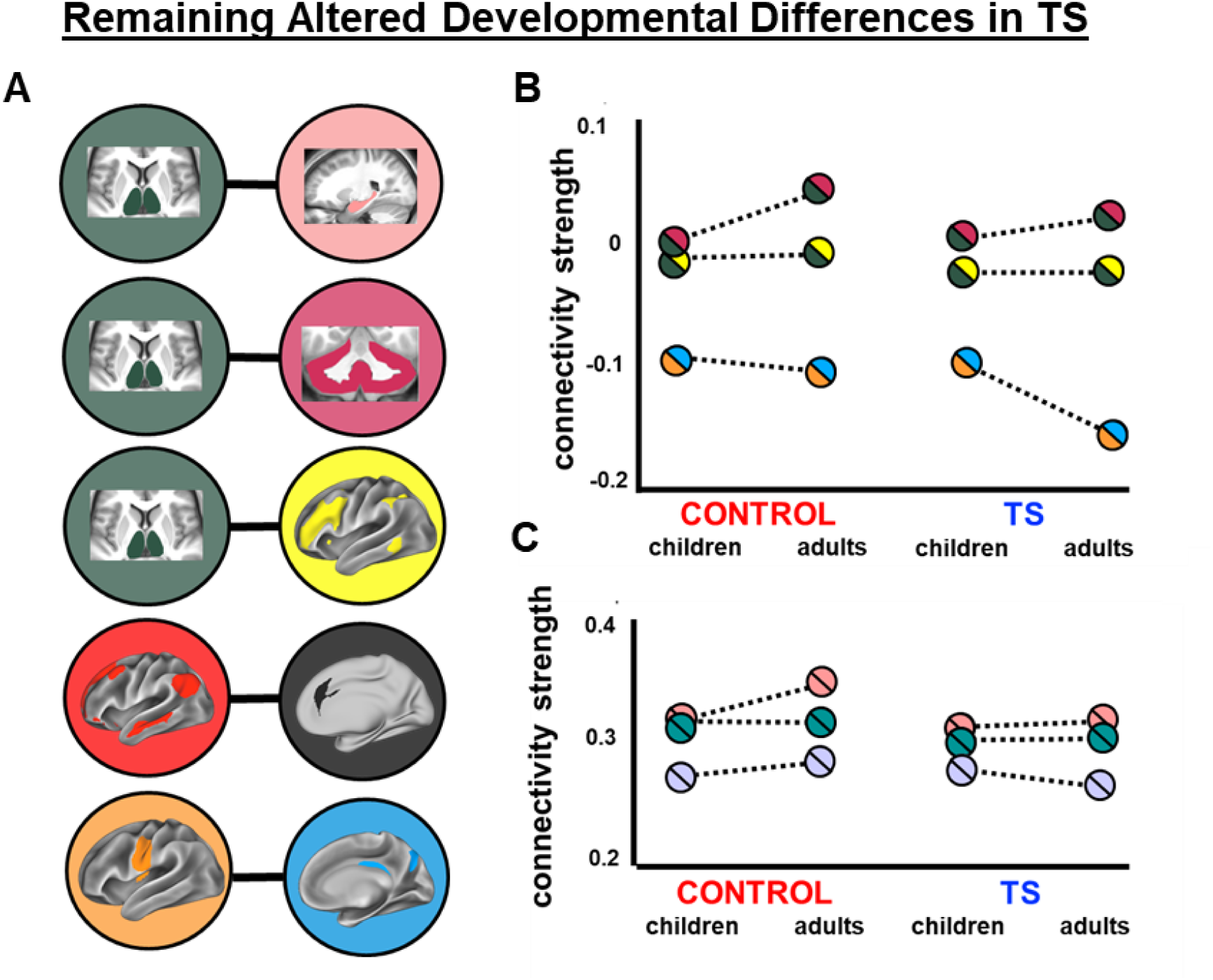
Blocks with altered developmental differences in TS that were not similar to other atypical development in TS.

